# White matter hyperintensities modify relationships between corticospinal tract damage and motor outcomes after stroke

**DOI:** 10.1101/2023.10.29.23297734

**Authors:** Jennifer K. Ferris, Bethany P. Lo, Giuseppe Barisano, Amy Brodtmann, Cathrin M. Buetefisch, Adriana B. Conforto, Miranda H. Donnelly, Natalia Egorova-Brumley, Kathryn S. Hayward, Mohamed Salah Khlif, Kate P. Revill, Artemis Zavaliangos-Petropulu, Lara A. Boyd, Sook-Lei Liew

## Abstract

Motor outcomes after stroke relate to corticospinal tract (CST) damage. Concurrent damage from white matter hyperintensities (WMHs) might impact neurological capacity for recovery after CST injury. Here, we evaluated if WMHs modulate the relationship between CST damage and post-stroke motor impairment outcome.

We included 223 individuals from the ENIGMA Stroke Recovery Working Group. CST damage was indexed with weighted CST lesion load (CST-LL). Mixed effects beta-regression models were fit to test the impact of CST-LL, WMH volume, and their interaction on motor impairment.

WMH volume related to motor impairment above and beyond CST-LL (β = 0.178, p = 0.022). We tested if relationships varied by WMH severity (mild vs. moderate-severe). In individuals with mild WMHs, motor impairment related to CST-LL (β = 0.888, p < 0.001) with a CST-LL x WMH interaction (β = -0.211, 0.026). In individuals with moderate-severe WMHs, motor impairment related to WMH volume (β = 0.299, p = 0.044), but did not significantly relate to CST-LL or a CST-LL x WMH interaction.

WMH-related damage may be under-recognised in stroke research as a factor contributing to variability in motor outcomes. Our findings emphasize the importance of brain structural reserve in motor outcomes after brain injury.

## Introduction

Upper-extremity motor impairment is one of the most common consequences of stroke (Lawrence *et al*., 2001) and typically results in long-term disability. (Meyer *et al*., 2015) The degree of damage to the corticospinal tract (CST) relates strongly to motor impairment after stroke, (Feng *et al*., 2015; Boyd *et al*., 2017) indicating a primary insult to the motor system. However, motor recovery after stroke is variable even after accounting for CST damage (Hayward *et al*., 2017). Recovery after stroke is likely mediated by compensation of surviving neural substrate. (Murphy and Corbett, 2009) This suggests that the integrity of structures beyond the CST might be prognostic of motor recovery, (Plow *et al*., 2015; Hayward *et al*., 2022) as overall brain health may be important in explaining why two individuals with similar stroke lesions can experience very different trajectories of recovery. (Liew *et al*., 2023)

White matter hyperintensities (WMHs) of presumed vascular origin are the most common form of age-related cerebrovascular damage (Duering *et al*., 2023). They are present in more than half of people above the age of 60. (de Leeuw *et al*., 2001) Individuals with WMHs are more likely to experience a stroke (Debette and Markus, 2010), in part due to common cardiometabolic risk factors between WMHs and stroke. (Jeerakathil *et al*., 2004) There is growing evidence that WMHs can also impact functional outcomes after stroke. (Georgakis *et al*., 2019) The relationship between WMHs and post-stroke cognitive impairment has been well established, (Georgakis *et al*., 2019) however, there have been few investigations of the specific impact of WMHs on motor outcomes after stroke. WMHs modulate relationships between stroke lesion volume and overall functional outcome. (Helenius and Henninger, 2015; Patti *et al*., 2016) Motor outcomes after stroke may similarly be modulated by concurrent WMHs, because of the widespread impacts of WMHs on cerebral networks, (Kim *et al*., 2015; Tuladhar *et al*., 2020) which may create pre-existing damage in compensatory pathways and therefore decrease the brain’s capacity for motor recovery.

Here, we tested if the relationship between post-stroke motor impairment and CST damage is impacted by concurrent WMH damage, controlling for age, time post-stroke and stroke lesion volume. Our study comprised a cross-sectional cohort of individuals in the subacute and chronic phases of recovery post-stroke. CST damage was operationalized by CST lesion load (CST-LL), an index of the degree of overlap between the CST and the stroke lesion. (Feng *et al*., 2015) CST-LL is a biomarker of post-stroke motor outcomes. (Boyd *et al*., 2017) WMH damage was operationalized by whole-brain WMH volume, measured with automated WMH segmentation. (Ferris *et al*., 2023)

We hypothesized that the relationship between motor impairment and CST-LL would be attenuated in individuals with higher WMH volumes, indicating a greater influence of concurrent WMHs on motor outcomes after stroke. This hypothesis was tested over two analyses: 1) a beta regression analysis testing the effects of WMH volume, CST-LL, and their interaction on motor impairment across the whole sample, and 2) beta regression analyses in subsamples stratified by severity of WMHs (mild versus moderate-severe WMHs) to assess if WMH severity modifies relationships between motor impairment, CST-LL, and WMH volume.

## Methods

### Study data

This study used cross-sectional multi-site data from the ENIGMA Stroke Recovery Working Group. (Liew *et al*., 2022) Data for this analysis were frozen on March 17^th^, 2023. The inclusion criteria for this analysis were: 1) individuals with stroke where imaging and behavioural assessments occurred >7 days post-stroke; 2) availability of T1-weighted MRI scans to index stroke lesions and either FLAIR or T2-weighted scans to index WMHs; 3) availability of a sensorimotor outcome measure (e.g., Fugl-Meyer Assessment, Action Research Arm Test, Wolf Motor Function Test). We extracted the following demographic information: age, sex, and days post-stroke. Stroke chronicity was defined as: early subacute phase of recovery >7 and ≤ 90 days post-stroke, late subacute phase > 90 and ≤180 days post-stroke, and chronic phase >180 days post-stroke. (Bernhardt *et al*., 2017)

Ethics approval was obtained from the local research ethics board of each contributing site. Written informed consent was obtained from all study participants in accordance with the Declaration of Helsinki. This study protocol was reviewed and approved by members of the ENIGMA Stroke Recovery Working Group. Data produced in the present study are available upon reasonable request to the corresponding author.

### Motor impairment score

Sensorimotor outcome measures were harmonized across study cohorts with a motor outcome score, consistent with previous ENIGMA publications. (Liew *et al*., 2021, 2023) The harmonized motor outcome score is a proportion of the total possible score for each sensorimotor scale. In the current analysis, 0% indicates no sensorimotor impairment and 100% indicates maximum sensorimotor impairment. For simplicity, we refer to this harmonized score as “motor impairment”.

### Lesion analysis

MRI processing was conducted with tools from FSL (v.6.0.5) and Freesurfer (v.7.2). Stroke lesions were manually traced on T1-weighted scans by trained researchers following established ENIGMA lesion tracing protocols. (Liew *et al*., 2018; Lo *et al*., 2023) WMHs were segmented with Freesurfer’s SAMSEG, which we previously established has robust performance in multi-site data from individuals with stroke. (Ferris *et al*., 2023) Linearly co-registered T1 and FLAIR/T2 scans were used as inputs into SAMSEG, and a 0.1 probability threshold was applied to tissue segmentation. We subtracted stroke lesion masks from segmented WMHs to prevent any misclassification of stroke lesions as WMHs. Fazekas scores were visually rated for each scan by a neuroradiologist. (Fazekas *et al*., 1987)

We calculated weighted CST-LL from stroke lesion masks. T1 images were non-linearly registered to MNI152 space using FSL’s FNIRT. (Andersson *et al*., 2007) To improve non-linear registration, we performed enantiomorphic normalization of the stroke lesion-by filling the stroke lesion with a copy of intact cerebral tissue from the opposite cerebral hemisphere. (Nachev *et al*., 2008; Ferris *et al*., 2022) WMH masks were incorporated as a cost-function mask, down-weighting their influence on non-linear registration. (Ferris *et al*., 2022) Stroke lesion masks were transformed to atlas space and overlaid onto CST derived from the JHU white matter atlas. Weighted CST-LL was calculated as a sum of the cross-sectional area of overlap between the stroke lesion and the CST, weighted by the maximum cross-sectional area of the CST. (Feng *et al*., 2015) This metric quantifies the degree of injury to the CST and accounts for the narrowing of the CST at the internal capsule, with higher CST-LL indicating more extensive overlap of the stroke lesion with the CST.

### Statistical analysis

Statistical analyses were conducted in R (v.4.3.1). For our primary analysis, motor impairment was modelled with beta regression. Beta regression was well-suited to our data because motor impairment scores were heavily right skewed and are a bounded proportion (from [0, 1]). (Ferrari and Cribari-Neto, 2004; Swearingen *et al*., 2011) We fit mixed-effects beta regressions with the ‘glmmTMB’ package. (Brooks *et al*., 2017) We tested the effects of WMH volume, CST-LL, and their interaction on motor impairment, with age, days post-stroke, and whole-brain stroke lesion volume as covariates and random slopes to control for study site. In line with recommendations from the Canadian Institutes for Health Research Sex and Gender-based Analysis policy (Canadian Institutes of Health Research, 2021), we conducted a supplementary analysis to explore if relationships with motor impairment varied by sex. In this supplementary analysis we included sex as an interaction term with CST-LL and WMH volume.

We tested if relationships with motor impairment varied by the severity of WMHs. We stratified the sample into “mild” and “moderate-severe” WMH subgroups with a cut-off for moderate-severe of ≥10mL WMH volume. This cut-off corresponds to a moderate Fazekas severity rating (periventricular or deep WMH Fazekas score of 2) (Joo *et al*., 2022) and is a posited threshold for WMH-related neurological changes. (DeCarli *et al*., 1995) We ran beta regression models within each subgroup. The subgroups were underpowered to fit random effects by site, therefore these beta regression models were fit without random effects.

## Results

Data from 223 individuals with stroke (82 females, 141 males) from 7 contributing research sites across four countries met our study inclusion criteria (median (IQR) age: 67 (58, 75); days post-stroke: 147 (92, 1,300); motor impairment: 5% (0%, 27%); stroke volume (mL): 2.2 (0.6, 16.5); WMH volume (mL): 5.3 (1.8, 11.7)). In terms of chronicity, 24% of the sample (n = 54) were in the early subacute phase of recovery (>7 and ≤ 90 days post-stroke), 27% (n = 60) were in the late subacute phase (> 90 and ≤180 days post-stroke) and 49% (n = 109) were in the chronic phase of recovery (>180 days post-stroke). (Bernhardt *et al*., 2017) See Supplementary Table 1 for a summary of participant characteristics by research site.

WMH volumes were significantly related to Fazekas scores (β = 0.106, p < 0.001; see Supplementary Figure 1), indicating that SAMSEG accurately estimated WMH volumes.

### Higher CST-LL and WMH volume relate to worse motor impairment after stroke

We tested our hypothesis that CST-LL, WMH volume, and their interaction relate to motor impairment with mixed effects beta regression, controlling for age, days post-stroke, stroke lesion volume, and research site (as a random effect). Greater CST-LL and larger WMH volumes were related to more severe motor impairment, holding all other factors constant (CST-LL: β = 0.812, p < 0.001; WMH volume: β = 0.178, p = 0.022; Table 1, Figure 1). Age, days post-stroke, and stroke volume also significantly related to motor impairment (See Table 1). The interaction term between CST-LL and WMH volume was not significant (β = -0.115, p = 0.265). There was no significant interaction effects with sex, indicating that relationships between motor impairment and CST-LL or WMH volume did not vary by sex (see: Supplementary Table 2).

**Table 1:** Relationships between motor impairment and stroke/WMHs.

| Predictor | Estimate | Std error | p value |
| --- | --- | --- | --- |
| Fixed |  |  |  |
| Age | -0.332 | 0.076 | **<0.001** |
| Days post-stroke | 0.335 | 0.084 | **<0.001** |
| Stroke volume | -0.228 | 0.103 | **0.027** |
| CST lesion load | 0.812 | 0.241 | **0.001** |
| WMH volume | 0.178 | 0.078 | **0.022** |
| CST lesion load:WMH volume | -0.115 | 0.103 | 0.265 |
| Random |  |  |  |
| $\sigma^2$ | 0.469 | | |
| $N_{site}$ | 7 | | |

Summary statistics and standardized parameter estimates from mixed-effects beta regression models, with harmonized motor impairment score as the outcome measure. Bold values indicate statistical significance ( $p < 0.05$ ). Abbreviations: CST = Corticospinal tract, WMH = White matter hyperintensity

**Figure 1:**
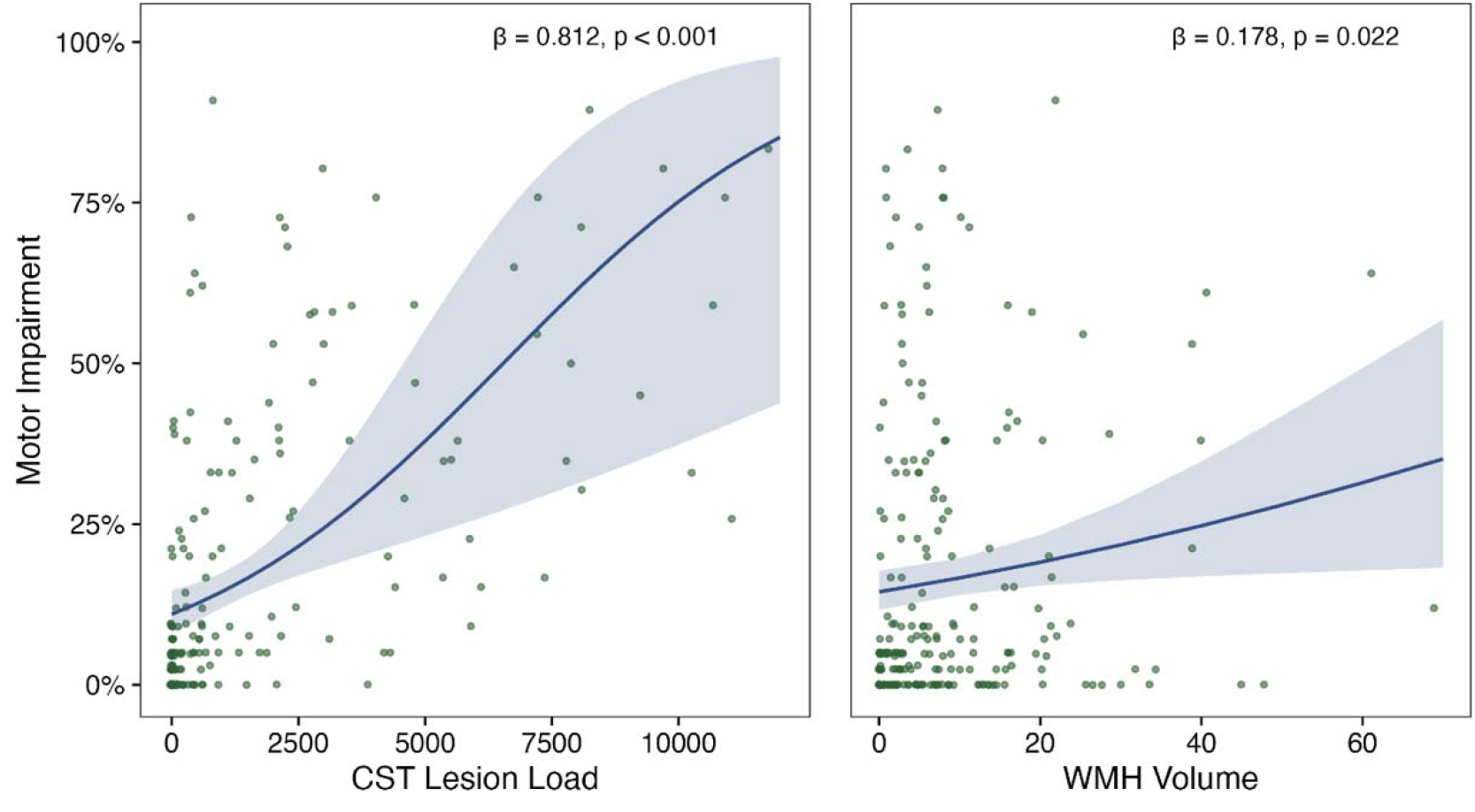
Relationships between motor impairment and stroke/WMHs Motor impairment by CST Lesion Load (left panel) or WMH volumes (in mL; right panel). Plots present beta regression line (solid),standard error (shaded), and parameter estimates (text)

### WMH severity modifies relationships between motor impairment and CST-LL/WMHs

There were 162 individuals with mild WMHs and 61 individuals with moderate-severe WMHs in our sample. Wilcoxon rank sum tests revealed that individuals with mild WMHs were younger than individuals with moderate-severe WMHs (W = 2824, p < 0.001), but there were no differences between groups in CST-LL (W = 4805, p = 0.748), stroke lesion volume (W = 5097, p = 0.717), or severity of motor impairment (W = 4729, p = 0.618) (see Supplementary Figure 2).

In individuals with mild WMHs, motor impairment was significantly related to CST-LL (β = 0.888, p < 0.001), with a significant CST-LL x WMH volume interaction (β = -0.211, 0.026) indicating individuals with smaller WMH volumes had a stronger relationship between CST-LL and motor impairment (see Table 2 and Supplementary Figure 3). In individuals with moderate-severe WMHs, motor impairment related to WMH volume (β = 0.299, p = 0.044), but not CST-LL (β = 0. 0.332, p = 0.120), with no significant CST-LL x WMH volume interaction (see Table 2). Figure 2 plots relationships between motor impairment and CST-LL for each WMH severity subgroup.

**Table 2:** Relationships between motor impairment and stroke/WMHs, stratified by WMH severity.

| Predictor | Mild WMHs |  |  | Moderate-Severe WMHs |  |  |
| --- | --- | --- | --- | --- | --- | --- |
|  | Estimate | Std error | p value | Estimate | Std error | p value |
| Age | -0.296 | 0.084 | **<0.001** | -0.278 | 0.153 | 0.070 |
| Days post-stroke | 0.405 | 0.090 | **<0.001** | 0.567 | 0.165 | **0.001** |
| Stroke volume | -0.194 | 0.111 | 0.081 | -0.088 | 0.184 | 0.633 |
| CST lesion load | 0.888 | 0.145 | **<0.001** | 0.332 | 0.213 | 0.120 |
| WMH volume | 0.057 | 0.086 | 0.503 | 0.299 | 0.149 | **0.044** |
| CST lesion load:WMH volume | -0.211 | 0.095 | **0.026** | 0.079 | 0.200 | 0.692 |

Summary statistics and standardized parameter estimates from beta regression models, with harmonized motor impairment score as the outcome measure. Bold values indicate statistical significance ( $p < 0.05$ ).
Abbreviations: CST = Corticospinal tract, WMH = White matter hyperintensity

**Figure 2:**
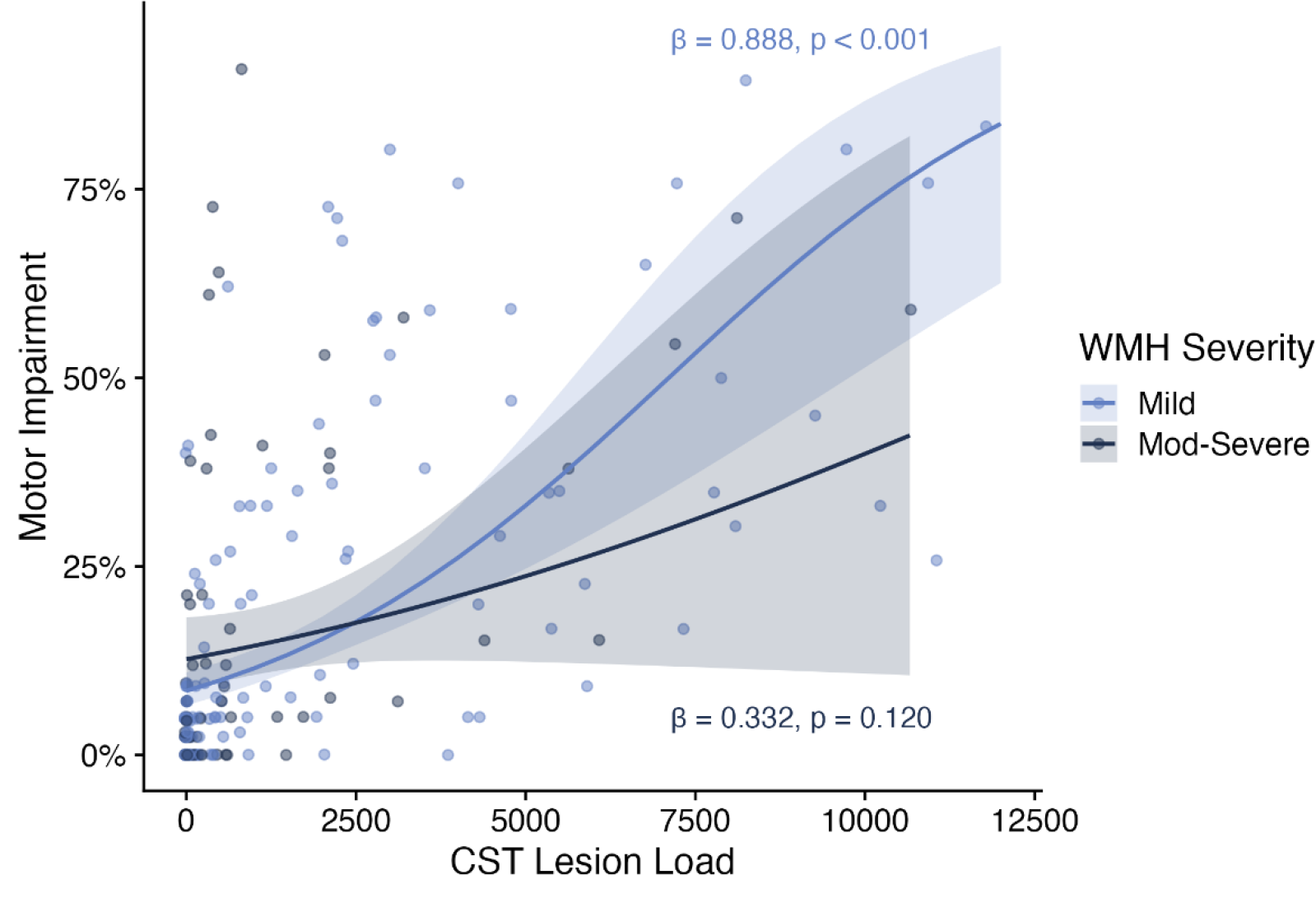
Relationships between motor impairment and CST Lesion Load stratified by WMH severity Motor impairment by CST Lesion Load, stratified by WMH severity (light blue = mild WMHs, dark blue = moderate-severe WMHs). Plots present beta regression line (solid), standard error (shaded), and parameter estimates (text) for stratified models

## Discussion

In this study, WMH volume related to post-stroke motor impairment over and above CST-LL and stroke volume. WMH severity was an effect modifier (Corraini *et al*., 2019) of CST-LL and motor impairment relationships, meaning the relationship between motor impairment and CST-LL varies in subgroups stratified by WMH severity. In individuals with mild WMHs there was a significant interaction between CST-LL and WMH volume, such that the relationship between motor impairment and CST-LL was attenuated with larger WMH volumes. In individuals with moderate-severe WMHs, motor impairment related to WMH volume and did not significantly relate to CST-LL. Our findings provide preliminary cross-sectional evidence that WMHs may be an important consideration in building prognostic neurological models of stroke recovery, especially for individuals with existing moderate to severe WMHs.

It is important to contextualize our findings with the existing literature on the CST damage and post-stroke motor outcomes. First, regardless of statistical significance, the effect sizes of CST-LL parameter estimates were greater than those for WMH volume across all tested models. This underscores the importance of CST damage as an explanatory variable of motor impairment after stroke (see consensus statement from Boyd *et. al*., 2017 (Boyd *et al*., 2017)). Secondly, our mild and moderate-severe WMH groups did not vary in severity of motor impairment. Therefore, it was not the case that individuals with moderate-severe WMHs (exceeding 10mL) had worse motor outcomes than individuals with mild WMHs. Rather, we saw that with increased WMH severity, WMH explained greater variability in motor impairment and CST-LL explained less variability in motor impairment. This suggests that WMH severity is an effect modifier of lesion-behaviour relationships, not an interactive factor. (Corraini *et al*., 2019) In other words, while WMH volume related to motor impairment across the whole sample, CST-LL and WMH did not have joint synergistic effects on motor impairment. Instead, individuals with moderate-severe WMHs may represent a neurologic subgroup where concurrent age-related cerebrovascular damage has greater explanatory power in post-stroke motor outcomes.

Very few studies to date have examined the impact of WMHs on motor systems, and research on the impact of WMHs on post-stroke motor outcomes has been equivocal. WMH volume in chronic stroke related to Wolf Motor Function Score in two previous reports. (Hicks *et al*., 2018; Auriat *et al*., 2019) WMH severity in acute stroke consistently relates to total Functional Independence measure (FIM) score, but the motor subscale of FIM related to WMH severity in some reports, (Senda *et al*., 2016) but not in others. (Hawe *et al*., 2018; Khan *et al*., 2019) Our study is the first to consider the combined effects of WMHs and CST damage on post-stroke motor outcome. Our results align with the work of Helenius and Henninger, (Helenius and Henninger, 2015) who found that individuals with moderate-severe WMHs had an attenuated relationship between stroke lesion volume and overall stroke severity as measured by the NIHSS.

The current study contributes important cross-sectional evidence for the impact of WMHs on motor outcomes after stroke. Future research should extend upon the current findings using longitudinal designs to test the effects of concurrent WMHs on trajectories of motor recovery. The strengths of our study are the large heterogeneous sample of individuals with stroke, and the use of robust methodologies to extract candidate neuroimaging predictors. Though data for this study came from different sites, manual stroke lesion drawings were performed at a single center with standardized protocols. (Lo *et al*., 2023) Moreover, we have previously demonstrated that our automated WMH segmentation protocol is robust across multiple sites in individuals with stroke. (Ferris *et al*., 2023) A limitation of the multisite and secondary nature of our study sample is that we had limited availability of additional covariates that may influence WMH severity and stroke outcomes, most notably cardiometabolic risk factors such as diabetes and hypertension. (Jeerakathil *et al*., 2004) Another limitation is that our sample had mild motor impairment overall (median impairment of 5%), which is typical of neuroimaging samples of motor impairment after stroke. However we were limited in ability to test for specific imaging markers of severe upper-extremity impairment, (Hodics *et al*., 2006) a patient subgroup where neuroimaging biomarkers may have the greatest benefit for prognostication of recovery. (Hayward *et al*., 2022)

In conclusion, our results suggest that WMHs are an under-recognised factor in stroke motor recovery research. WMHs explained variability in motor impairment over and above stroke lesion volume and CST damage. Furthermore, WMH severity might define neurologic subtypes, wherein structural brain reserve has more explanatory power and CST damage has less explanatory power for individuals with extensive pre-existing damage to cerebral white matter. The structural reserve of the brain before a stroke injury is increasingly recognised as an important element predicting capacity for motor recovery. (Liew *et al*., 2023) Including WMHs in motor recovery research could advance models of neurological recovery by accounting for the full spectrum of cerebrovascular damage in the brain.

## Data Availability

Data produced in the present study are available upon reasonable request to the corresponding author.

