## Supplementary materials for "White matter hyperintensities modify relationships between corticospinal tract damage and motor outcomes after stroke"

| Supplementary Table 1: Participant demographics across contributing research sites | | | | | | | |
| --- | --- | --- | --- | --- | --- | --- | --- |
| **Characteristic** | **R024**, N = 21^1^ | **R041**, N = 114^1^ | **R042**, N = 34^1^ | **R045**, N = 4^1^ | **R046**, N = 4^1^ | **R048**, N = 29^1^ | **R057**, N = 17^1^ |
| Age | 63 (55, 68) | 70 (63, 78) | 48 (44, 60) | 62 (58, 64) | 66 (53, 76) | 71 (62, 74) | 65 (60, 72) |
| Sex |  |  |  |  |  |  |  |
| F | 11 (52%) | 38 (33%) | 18 (53%) | 1 (25%) | 1 (25%) | 10 (34%) | 3 (18%) |
| M | 10 (48%) | 76 (67%) | 16 (47%) | 3 (75%) | 3 (75%) | 19 (66%) | 14 (82%) |
| Days post stroke | 519 (305, 1,312) | 92 (84, 108) | 740 (533, 1,302) | 2,882 (2,211, 3,980) | 2,538 (1,389, 3,952) | 2,604 (1,568, 4,242) | 1,701 (592, 2,554) |
| Chronicity |  |  |  |  |  |  |  |
| Early subacute | 0 (0%) | 53 (46%) | 1 (2.9%) | 0 (0%) | 0 (0%) | 0 (0%) | 0 (0%) |
| Late subacute | 0 (0%) | 60 (53%) | 0 (0%) | 0 (0%) | 0 (0%) | 0 (0%) | 0 (0%) |
| Chronic | 21 (100%) | 1 (0.9%) | 33 (97%) | 4 (100%) | 4 (100%) | 29 (100%) | 17 (100%) |
| Motor impairment | 5% (5%, 5%) | 0% (0%, 2%) | 38% (33%, 52%) | 51% (39%, 63%) | 33% (19%, 49%) | 12% (8%, 59%) | 23% (9%, 73%) |
| Stroke (mL) | 3.7 (0.8, 31.7) | 0.9 (0.3, 7.5) | 10.8 (3.5, 43.2) | 6.2 (3.5, 8.3) | 3.6 (3.1, 8.9) | 4.4 (1.6, 20.3) | 15.7 (1.6, 97.8) |
| WMH (mL) | 1.9 (0.1, 11.7) | 4.8 (1.3, 11.6) | 6.9 (4.5, 15.6) | 0.7 (0.6, 1.3) | 4.1 (2.4, 8.1) | 5.9 (4.0, 11.2) | 6.1 (3.7, 10.1) |
| Total Fazekas score | 2.00 (2.00, 2.00) | NA | 2.00 (1.00, 2.75) | 1.50 (0.75, 2.00) | 1.50 (0.75, 2.50) | 2.00 (2.00, 3.00) | NA |
| ^1^Median (IQR); n (%) | | | | | | | |

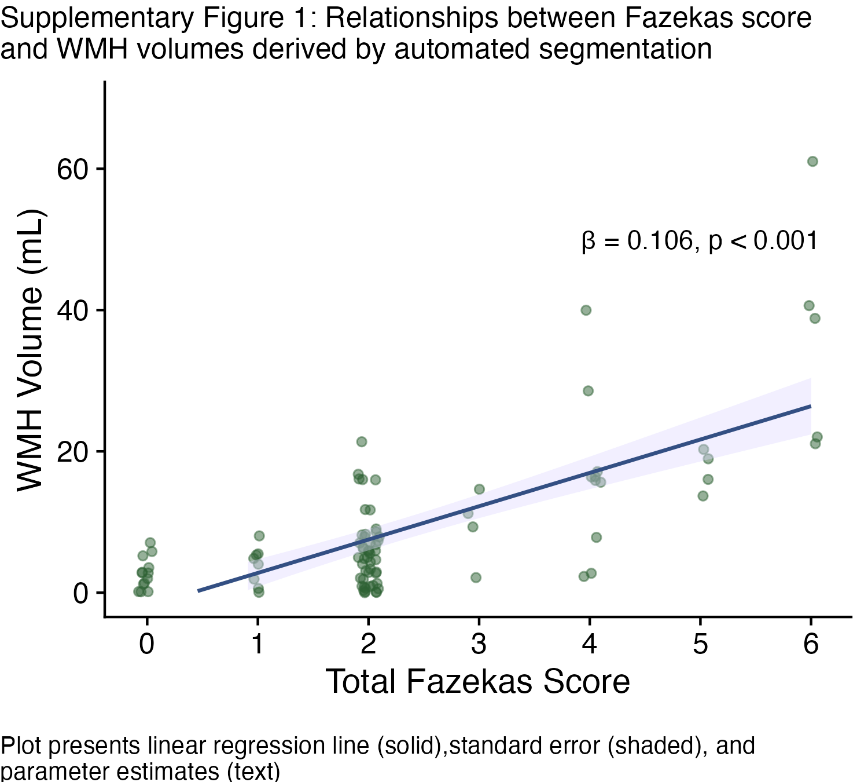

| Supplementary Table 2: Relationships between motor impairment and stroke/WMHs testing effects of sex | | | |
| --- | --- | --- | --- |
| Predictor | Estimate | Std error | p value |
| Fixed | | | |
| Age | -0.320 | 0.076 | **<0.001** |
| Days post-stroke | 0.349 | 0.084 | **<0.001** |
| Stroke volume | -0.218 | 0.104 | **0.036** |
| CST lesion load | 0.875 | 0.260 | **0.001** |
| WMH volume | 0.220 | 0.093 | **0.018** |
| Sex | -0.295 | 0.141 | **0.037** |
| CST lesion load:WMH volume | -0.077 | 0.129 | 0.551 |
| CST lesion load:Sex | -0.102 | 0.146 | 0.483 |
| WMH volume:Sex | -0.122 | 0.150 | 0.413 |
| CST lesion load:WMH volume:Sex | -0.083 | 0.227 | 0.715 |
| Random | | | |
| $\sigma^{2}$ | 0.486 |  |  |
| $N_{site}$ | 7 |  |  |
| Summary statistics and standardized parameter estimates from mixed-effects beta regression models, with harmonized motor impairment score as the outcome measure and males as the reference category for sex. Bold values indicate statistical significance (p < 0.05). Abbreviations: CST = Corticospinal tract, WMH = White matter hyperintensity | | | |

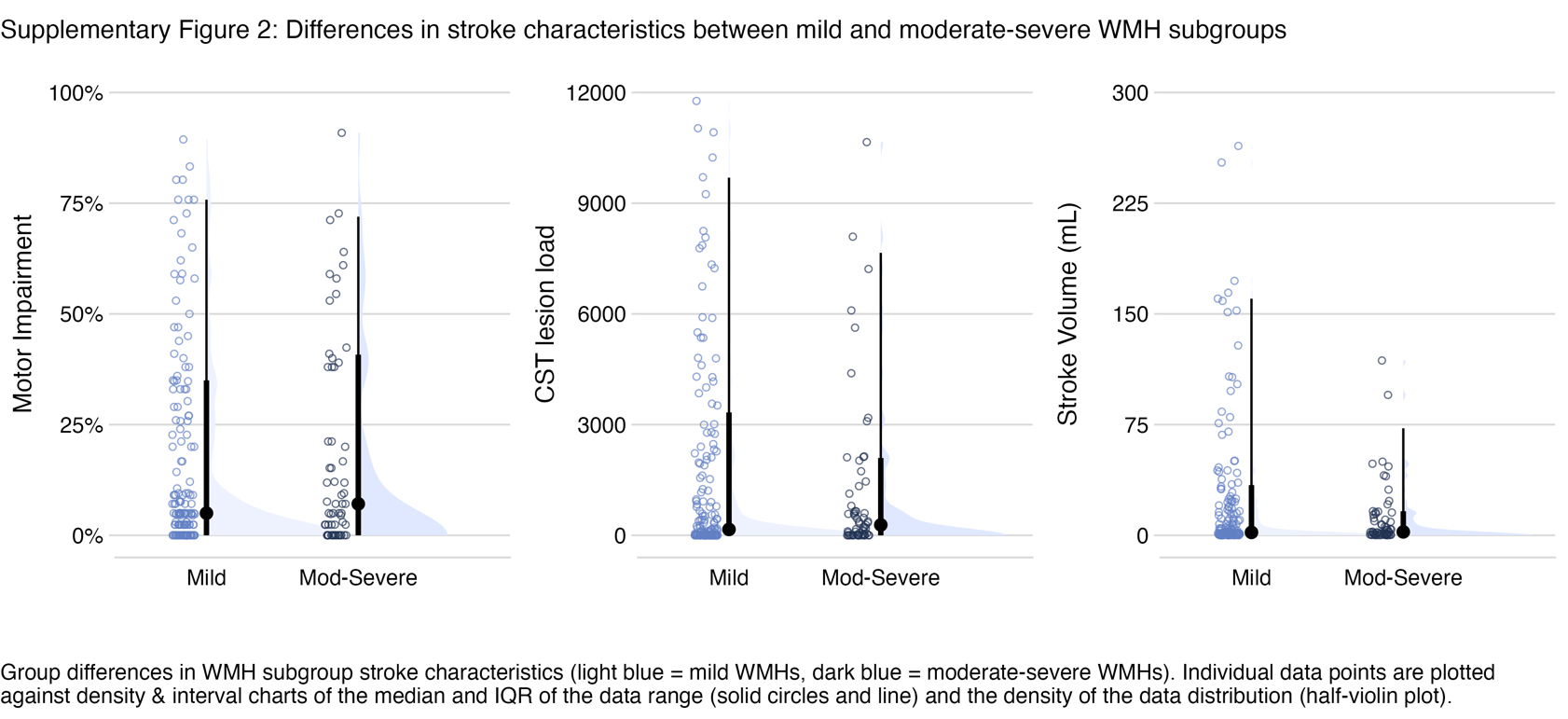

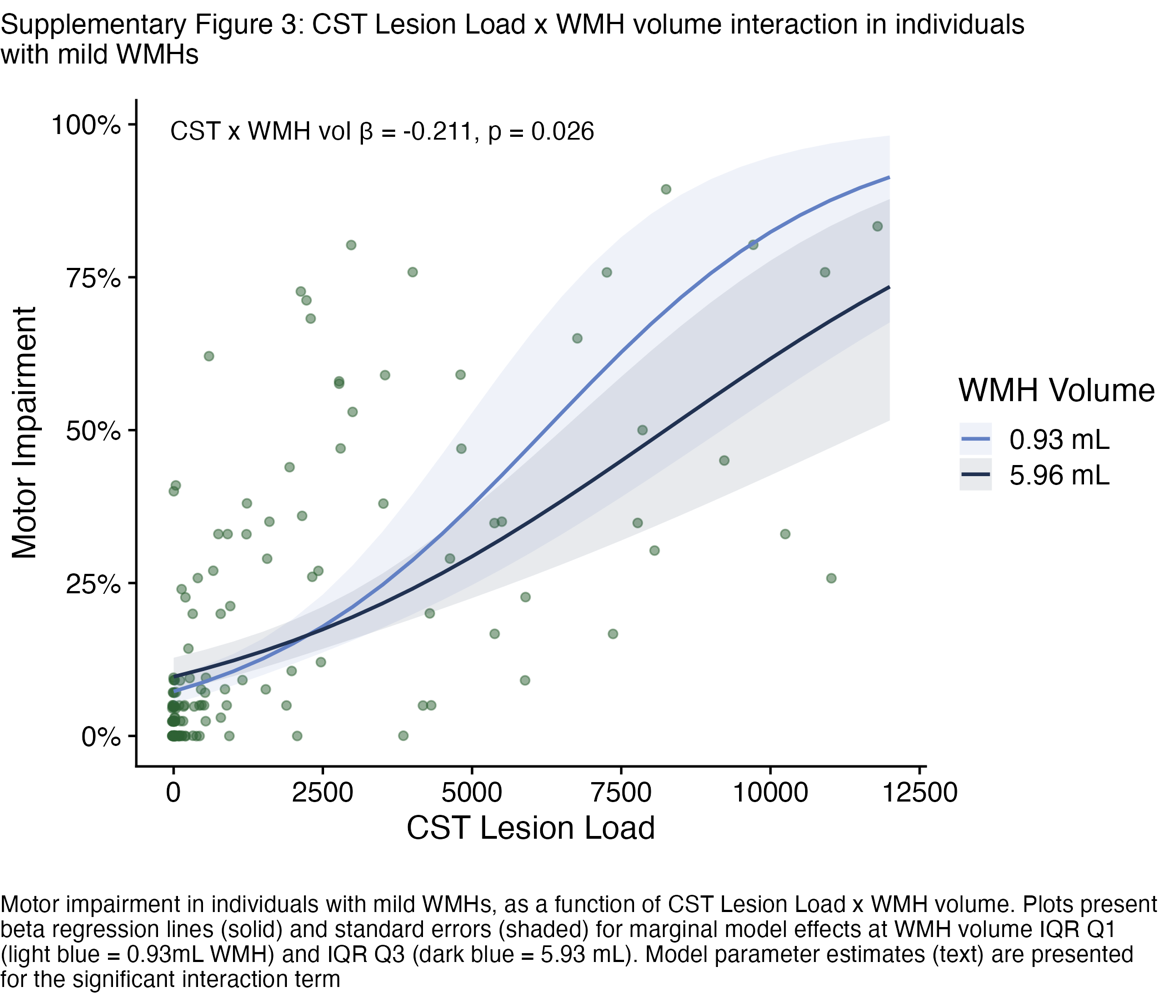
